# Reopening International Borders without Quarantine: Contact Tracing Integrated Policy against COVID-19

**DOI:** 10.1101/2021.06.10.21258672

**Authors:** Zidong Yu, Xiaolin Zhu, Xintao Liu, Xiang Chen, Tao Wei, Hsiang-Yu Yuan, Yang Xu, Rui Zhu, Huan He, Hui Wang, Man-sing Wong, Peng Jia, Wen-zhong Shi, Wu Chen

## Abstract

With the COVID-19 vaccination widely implemented in most countries, propelled by the need to revive the tourism economy, there is a growing prospect for relieving the social distancing regulation and reopening borders in tourism-oriented countries and regions. The need incentivizes stakeholders to develop border control strategies that fully evaluate health risks if mandatory quarantines are lifted. In this study, we have employed a computational approach to investigate the contact tracing integrated policy in different border reopening scenarios in Hong Kong, China. Built on a modified SEIR epidemic model with a 30% vaccination coverage, the results suggest that scenarios with digital contact tracing and quick isolation intervention can reduce the infectious population by 92.11% compared to those without contact tracing. By further restricting the inbound population with a 10,000 daily quota and applying moderate-to-strong community non-pharmacological interventions (NPIs), the average daily confirmed cases in the forecast period of 60 days can be well controlled at around 9 per day (95% CI: 7–12). Two main policy recommendations are drawn from the study. First, digital contact tracing would be an effective countermeasure for reducing local virus spread, especially when it is applied along with a moderate level of vaccination coverage. Second, implementing a daily quota on inbound travelers and restrictive community NPIs would further keep the local infection under control. This study offers scientific evidence and prospective guidance for developing and instituting plans to lift mandatory border control policies in preparing for the global economic recovery.

## Introduction

According to the World Tourism Organization (UNWTO), 2020 was the documented worst year for international tourism with an unprecedented recession of 74% (UNWTO, 2021). This economic downturn was caused by the global spread of the coronavirus disease 2019 (COVID-19), whereas many countries closed their borders to contain the virus spread resulted from international travel and trade. These border control strategies had a lasting impact on countries and regions relying on the tourism economy, such as France, Thailand, and Hong Kong, China (Yang et al., 2020). Further, the shortage of cross-border labor was predisposed to restrict the production capacity in certain industries, such as outsourced agriculture (Hobbs, 2021). As phases of COVID-19 vaccination are rolling out and new cases are declining in most world regions, growing discussion arises about when and how to lift international travel bans and reopen borders in the prospect of reviving the economy from the global calamity.

To minimize the risk for epidemic resurgence, some countries have partially reopened their borders by implementing strict quarantine rules on all incoming travelers. For example, Canada imposed mandatory nucleate tests as well as a 14-day quarantine policy (i.e., 3-day institutional quarantine and 11-day self-quarantine) on all incoming travelers. In Hong Kong, all incoming travelers were required to undergo a 14-day or 21-day mandatory quarantine at designated institutions, whereas the quarantine period was determined on the risk level of their country of origin. More importantly, the mandatory quarantine helped public health officials to isolate asymptomatic cases in the incubation period before the symptom onset. These strict containment measures were proven to be effective in reducing imported cases and their potential secondary local transmission (Chinazzi et al., 2020; Wells et al., 2020). Although the enforcement of the quarantine played an essential role in containing the virus spread, it was criticized for posing considerable obstacles for human movement and international trade (Dickens et al., 2020). As a result, stakeholders were pressed to reevaluate the health risk associated with the possibility of reopening borders without an elongated period of quarantines to offset the socioeconomic loss. Eventually, it is expected that the reopening policy will achieve an intricate balance between acceptable levels of health risks and steady socioeconomic recovery.

Contact tracing is a promising way to monitor the spread of infectious cases. It was demonstrated in clinical research and simulation results that contact tracing was highly effective in identifying the etiology of transmission and helping maintain low infection levels (Ferretti et al., 2020; Hellewell et al., 2020; Quach and Hoang, 2020). Given that a local spread can quickly induce a spike of new cases, conventional contact tracing methods, such as the travel diary survey, were characterized by a delay between confirming a new case and locating the patient’s close contacts (Peak et al., 2020). In contrast, digital contact tracing enabled by location-based devices (e.g., mobile phones, fitness tracking bands) make epidemiological investigation highly efficient and the follow-up notification process in near real-time upon the confirmation of new cases (Yang et al., 2021).

In addition to contact tracing, non-pharmaceutical interventions (NPIs) have been implemented in most countries to mitigate the spread of the disease. NPIs for domestic containment can be broadly categorized into two strategies—one focused on personal protection and the other on community interventions (Centers for Disease Control and Prevention, 2020). For the latter strategy, it was evidenced that community social distancing, such as school arrangement, work-from-home arrangements, and limited opening of non-essential businesses, were effective to lower the intensity of interpersonal interactions and thus alleviate the health impact before the vaccine became widely available (Davies et al., 2020; Flaxman et al., 2020; Kissler et al., 2020).

With the aim to reopen international borders, some researchers articulated the potential effects of the imported virus carriers and relieving NPIs (Dickens et al., 2020; Smith et al., 2021). However, these recommendations were mostly based on mandatory quarantines, and there has been limited scientific exploration to quantify the risk of COVID-19 if borders are reopened and mandatory quarantines are lifted. This oversight could be problematic since the pathway towards rehabilitating the travel normalcy must include a thorough understanding of the health risk if the quarantine process is to be circumvented for international travel.

To this end, there is an urgent need to formulate a restrictive but sustainable plan for reopening international borders by relieving the quarantine requirements and simultaneously applying digital contact tracing. In this paper, we propose a framework to integrate the importation risk induced by potential virus carriers into a mechanistic epidemic model in a case study of Hong Kong. We aim to answer two important research questions: (1) To what extent can digital contact tracing help contain the local infection compared to scenarios without contact tracing? (2) To what extent can the containment measures be improved, such as combining digital contact tracing with the inbound quota and community NPIs?

## Results

### Deriving parameters from empirical data using a modified SEIR model

To estimate the course of local infections, we have extracted confirmed cases in Hong Kong, which were further categorized as either local cases or cases linked to local cases from December 25, 2020 through May 5, 2021 (Fig. 1a). The classic process-based Susceptible-Exposed-Infectious-Removed (SEIR) model is modified to account for local factors affecting the transmission (Methods). These factors include the community NPIs and the regional COVID-19 vaccination program (Supplementary Table 1 and Supplementary Fig. 1).

**Fig. 1.**
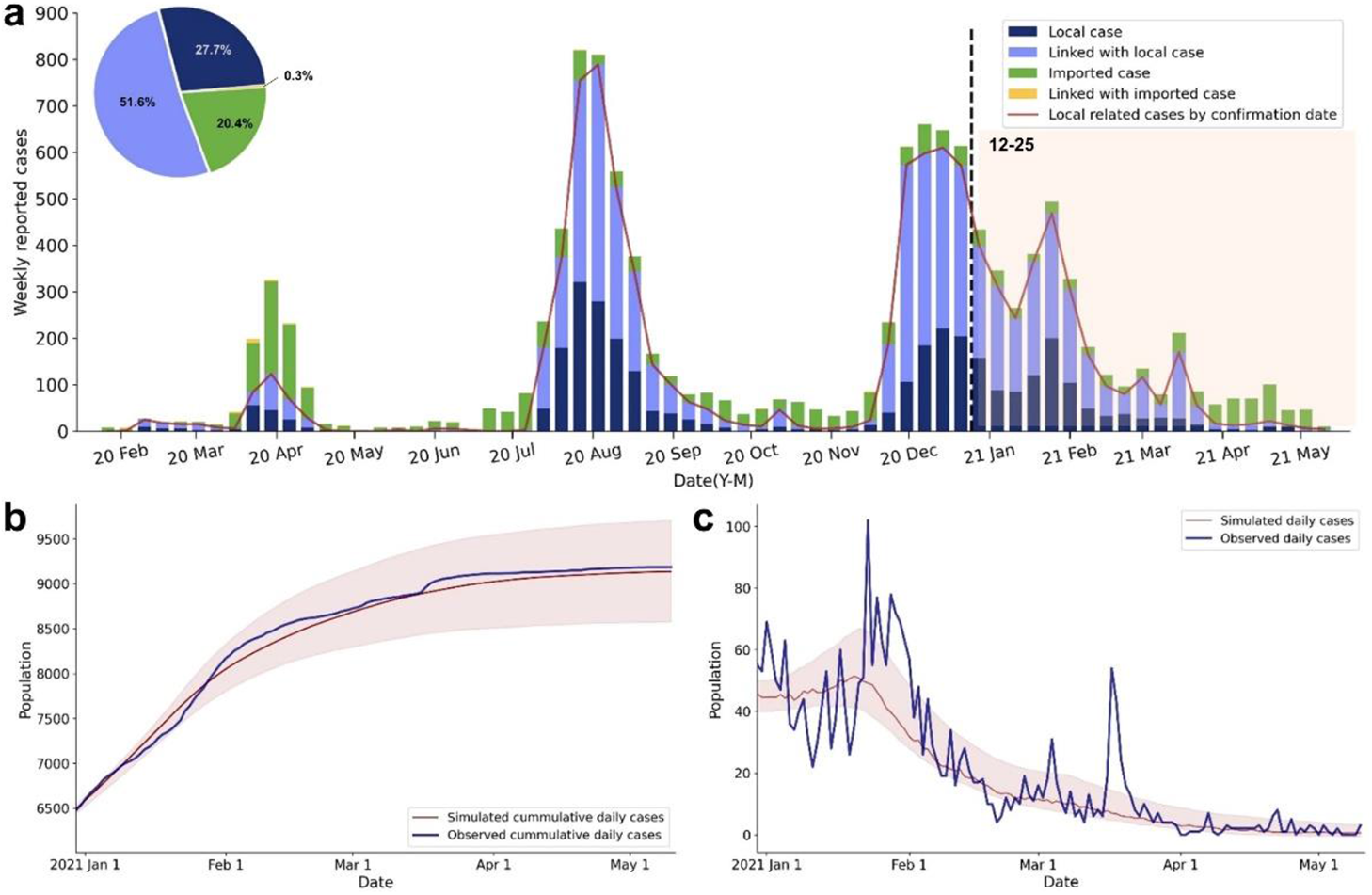
Reported cases and fitted curve of the SEIR model in Hong Kong. **A**. The epidemic curve by weekly reported cases that belong to different case categories: local cases, cases linked to local cases, imported cases, and cases linked to imported cases. The dashed line and color band indicate the period of observation for model fitting. **B**. Comparison of the observed and simulated cumulative daily cases. The shaded areas are the 95% CI of the simulated cases. **C**. Comparison of the observed and simulated daily cases. The shaded areas are the 95% CI of the simulated cases.

The modeling results show a relatively high level of correspondence with observed cumulative cases, confirming that our modification of the classical SEIR model can well preserve the dynamic course of local infections (R^2^ = 0.99, RMSE = 129.04, Fig. 1b). The simulated daily new cases, derived from the fitted model, are compared with observed daily cases (Fig. 1c). However, the predictive results are less precise for January and from March 12 through 14, of which some cases are beyond the 95% confidence intervals (CI). Such deviations could be attributed to the stochastic nature of the local outbreaks, such as the superspreading events (SSEs) in a low-prevalence context. SSEs can dramatically increase the confirmed incidences in a short period (Leung et al., 2021). Explicitly, one explanation of the inconsistency during mid-to-late January is the explosive growth of reported cases due to the massive untraceable SSEs in Yau Ma Tei and Jordan districts. Besides, our prediction underestimates the reported cases dated from March 12 through 14, which are mostly associated with the outbreak of the URSUS Fitness cluster. Despite these SSEs, the modified SEIR model has a satisfactory capacity to elucidate the course of local infections via a robust simulation process.

The fitted parameters are estimated from our model by relating to the daily transmission rate (Supplementary Fig. 2). Among these NPIs, compulsory testing is estimated to have the largest alleviating effect on reducing the disease transmissivity by 48.69% (95% CI: 30.29–51.93%) The results provide evidence that the interventions in the past few months were most effective in reducing the transmission rate.

### Simulating the effects of contact tracing on reopening borders

In this section, we analyze our simulation results for estimating the difference between two designated baseline scenarios during the potential border-reopening phase: one with digital contact tracing and the other without digital contact tracing. We assume 152,473 people per day will enter the region, which was the historical visitor data in 2019 (Tourism Commission, 2020). In addition, COVID-19 vaccination helps protect the susceptible population by activating an antibody (immunity) response. Hypothetical scenarios are proposed to understand whether digital contact tracing can help to contain the importation risks when only a proportion of local residents are vaccinated.

The effectiveness of using digital contact tracing is estimated by assuming the full quarantine (100%) upon case identification under three levels of vaccination coverage (Fig. 2 and Supplementary Fig. 4). A large difference regarding the simulated daily cases from active infectious patients can be clearly observed between all baseline scenarios with and without contact tracing. In particular, 68.23%, 91.07%, and 97.24% of the infectious population are predicted to be isolated under the scenarios with 70%, 50%, and 30% vaccination coverage, respectively. Another finding is that a higher vaccination coverage of the residents can more effectively contain the virus spread by reducing the susceptible population. The optimal scenario can be concluded from the results, where applying digital contact tracing on all inbound travelers and a 70% vaccination coverage rate can largely flatten the epi curve and maintain the infections on a relatively low level. In this optimal scenario, we estimate the daily reported cases to be 188 (95% CI: 175–210) and the cumulative reported cases to be 19163 (95% CI: 17393–21761) on the 60^th^ day of reopening the border. However, as of May 26, 2021 or 88 days since the initiation of the vaccination program, only 934,399 (12.5% of the total population) received complete vaccination, meaning that the actual vaccinated population was likely to be far below the theoretical threshold for reaching herd immunity (GovHK, 2021). As such, we simulate the effectiveness of digital contact tracing under the most pessimistic but more realistic scenario with 30% vaccination coverage. In such settings, the daily reported cases are predicted to be 218 (95% CI: 110–268), and the cumulative reported cases are 26530 (95% CI: 22103–31602) on the 60^th^ day of reopening the border.

**Fig. 2.**
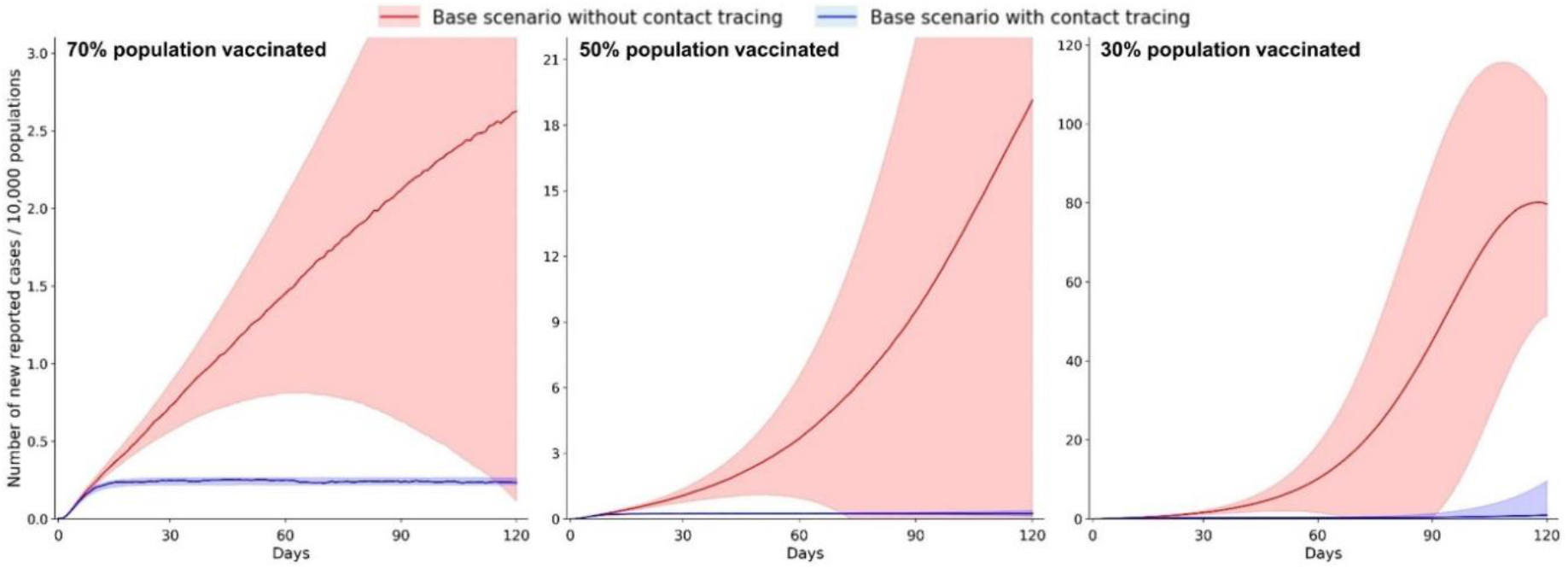
Estimated effectiveness of digital contact tracing in containing the importation risk for reopening borders during the 120-day simulation period. The daily reported cases are estimated under three different vaccination coverages (30%, 50%, and 70%). The shaded area shows the 95% CI of estimated daily reported cases.

### Joint effects of controlling arrival population and community NPIs

As shown above, the application of digital contact tracing can effectively contain the importation risk of COVID-19; however, it is less likely for public health officials to well control the transmission in the worst-case scenario (30% vaccination coverage). Hence, it is imperative to estimate the joint effects of controlling the inbound population and community NPIs. Specifically, we factor in the joint effects of applying a daily quota on inbound travelers and various degrees of community NPIs with and without contact tracing. Compared to the historical population flow (152,473 per day) in the baseline scenarios, two daily quotas are set: 76,236 (50% of historical arrivals) and 10,000. Additionally, moderate-to-strong community NPIs are applied, including the school arrangement, suspension of large events, public facility arrangement, restricting social gathering, using voluntary check-in system, and compulsory testing (Supplementary Table 2 and Supplementary Fig. 3).

As reported in our simulation results, the joint effects of controlling for inbound travelers and applying community NPIs are predicted to further reduce the number of daily infections compared to the scenario with contact tracing only (Fig. 3 and Supplementary Table 3). The alleviating effects are estimated to be more substantial with a limited daily quota and more restrictive community NPIs. Without implementing any community NPIs, the cumulative reported cases during the first 60 days after reopening the border are predicted to be 5008 (95% CI: 3859–7105) and 978 (95% CI: 151–2069) under moderate and strict border control policies, respectively. Similarly, when there is no daily quota, our simulation estimates that the cumulative reported cases are 9992 (95% CI: 9213–11150) and 10061 (95% CI: 9168–11119) under moderate and strong NPIs, respectively. Ideally, applying a 10,000 daily quota and moderate-to-strong community NPIs jointly can limit the cumulative reported cases to 549 (95% CI: 440–697), where daily reported cases is 9 (95% CI: 8–13) on the 60^th^ day of reopening the border.

**Fig. 3.**
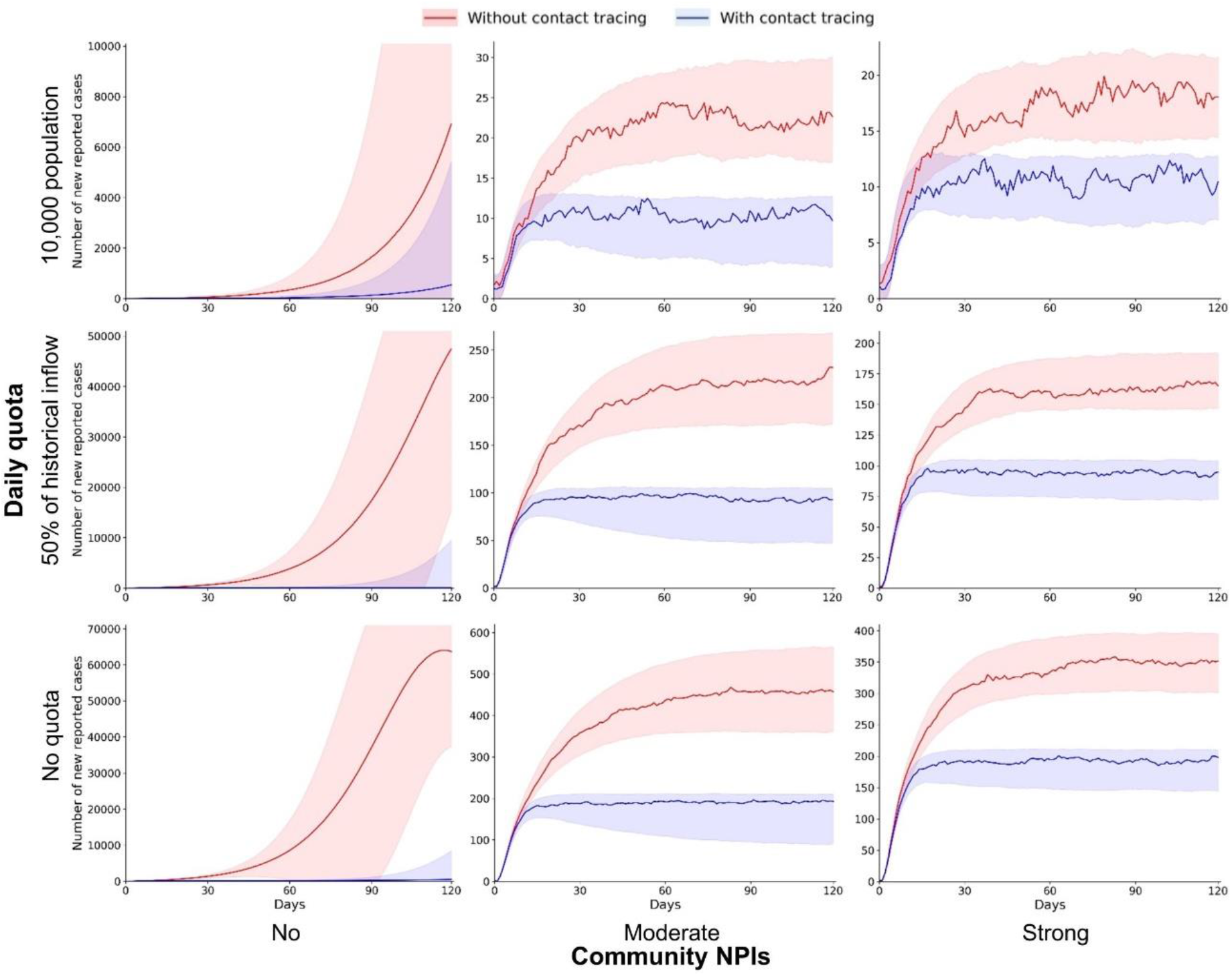
Implementing a daily quota for inbound travelers and community NPIs to contain the importation risk during the 120-day simulation period. The number of daily reported cases are estimated under scenarios with different daily quotas (no quota, 50% of historical inflow, and 10,000 population) and community NPIs (no, moderate, and strong). The shaded areas are the 95% CI of estimated daily reported cases.

In practical terms, digital contact tracing and follow-up intervention cannot be 100% successful for isolating infectious individuals. Hence, we further assume practical scenarios with a success rate of 90% for isolation (Supplementary Table 4 and Supplementary Fig. 5). Our simulation estimates that digital contact tracing with a 10,000 daily quota and moderate-to-strong community NPIs can still ensure the infections to be maintained on a low level, in which cumulative reported cases are limited to 634 (95% CI: 457–716) during the first 60 days of reopening the border. Overall, the results suggest that for cities with low vaccination coverage, restricting inbound travelers, applying community NPIs, and implementing digital contact tracing jointly can effectively contain the importation risk of COVID-19 while reopening borders.

## Discussion

Concerning the importation risk of COVID-19 during the border reopening phase, existing studies mainly articulated strategies about mandatory quarantines. However, a lengthy quarantine process could be unbearable for inbound travelers and can pose obstacles to the revival of the tourism economy. Using location-based devices, such as mobile phones and fitness tracking bands, to identify and track the close contacts of infectious patients has been proven successful in containing the virus spread (Altmann et al., 2020; Li and Guo, 2020). In this paper, we perform a simulation study to evaluate the effectiveness of digital contact tracing under different vaccination scenarios without mandatory quarantines. A modified stochastic SEIR model is built and applied to forecast the future development of the pandemic in Hong Kong.

By reconstructing the COVID-19 transmission from historical data, we confirm that digital contact tracing can effectively help to contain the importation risk of COVID-19 by flattening the epi curve. In the most realistic scenario with only a 30% vaccination rate, the predicted daily reported cases for the first 60^th^ days after reopening the border is 174 (95% CI: 146–207), which is a significant reduction of 92.11% (95% CI: 69.90 %–96.02%) compared to the setting without contact tracing. This finding provides evidence that digital contact tracing and the follow-up isolation process upon case confirmation can prevent the virus from further spreading and can thus reduce the transmissibility of the disease (Ferretti et al., 2020; Kucharski et al., 2020). Albeit the effectiveness of the digital contact tracing, additional containment measures are still needed to curb the pandemic. One unique feature of our model is to simulate the short-term stay of inbound travelers if the mandatory quarantine is relieved, which has been less mentioned by the discussion about reopening borders (Smith et al., 2021; Zhu et al., 2021). This feature can be further applied to modeling the outflow of asymptomatic travelers before their symptom onset in another country. This international application is highly relevant to studies of the importation risk around the globe because of the increasing international trend, travel, and migration when more borders are open.

Concurrent with digital contact tracing, restricting international travel and implementing community NPIs are recommended measures to contain the local infection. Explicitly, applying a daily quota on travelers can limit the likelihood of virus carriers entering the city, while community NPIs can further reduce the virus transmissibility. It is noteworthy that we have simulated the least restrictive but sustainable criteria for guiding the reopening policy (Haleem et al., 2020), where extreme containment measures, such as school closures, are not considered. We also simulated under the optimistic scenario with digital contact tracing, a daily quota on inbound travelers, and community NPIs jointly in effect, forecasting the daily increase in a 60-day period will be around 9 cases per day (95% CI: 7–12); and without contact tracing, the increase will be around 14 cases per day (95% CI: 12–17). However, as stressed by other studies, it is intractable to implement contact tracing with instantaneous isolation of infectious patients (Ivers and Weitzner, 2020; Kretzschmar et al., 2020). In this regard, the effectiveness of contact tracing in the optimistic scenario is highly reliant upon the response capacity of the local public health department. More significantly, while contract tracing relieves the workload and resources that could be otherwise expended on mandatory quarantines, it is expected that the potential surge of disease outbreaks can become a prime concern when there is no barrier to hold asymptomatic travelers.

Nevertheless, several limitations exist in our proposed framework and the designed scenarios. First, our simulation model is built on the epidemiological parameters derived from reported cases in Hong Kong. Based on recent COVID-19 studies and results of the Universal Community Testing Program, we hypothesize that all of the asymptomatic and symptomatic infectious patients are identified, diagnosed, and reported by the case surveillance system after the third-wave of outbreak in Hong Kong (Cao et al., 2020; The Government of the Hong Kong Special Administrative Region, 2020; To et al., 2020). Nevertheless, this assumption may not be applied to other countries and regions due to the heterogeneity in case diagnosis and surveillance, whereas the actual infections are estimated to be much greater than the reported cases, for example, 6–24 times higher in the United States (Allen et al., 2020; Havers et al., 2020). Second, real-time NPIs in the past few months are recorded and simulated in our model to understand their effectiveness. However, preventive behaviors on the individual level (e.g., human mobility, face covering), which dictate the likelihood of infection (Chan et al., 2021; Cowling et al., 2020), are not considered in our simulation. Thus, future extensions of the model should emphasize these behavioral factors to better estimate virus transmissibility. Third, our model can simulate the epi curve over a short period (e.g., two months) but is unable to predict results over the long term because of the many uncertainties arising from the epidemic development, such as the variants of the virus and the efficacy of vaccination (Chen et al., 2021; Vaidyanathan, 2021). These changing situations can be accommodated by learning and updating epidemiological parameters as the simulation progresses on a daily basis. Finally, it should be noted that there are ethical issues in the usage of digital contact tracing. Its wide implementation must be grounded on public trust and health policy about privacy protection (Ivers and Weitzner, 2020). These considerations will eventually help the public weigh the health benefits of digital contact tracing against the potential privacy issues.

In this research, we investigate the importation risk of COVID-19 in scenarios of reopening borders. This endeavor is an essential step to recover the economy in the post-pandemic phase. Two policy-relevant recommendations are drawn from our simulations. First, our results support that digital contact tracing is an effective countermeasure to limit case reproduction, especially when the imported case can be quickly identified and isolated. Second, restricting inbound travelers and implementing community NPIs jointly can ensure the local infection to be under control. Overall, this study justifies the effectiveness of digital contact tracing integrated policies in reducing the importation risk of COVID-19 and can offer an insight into sustainable border-reopening policies.

## Methods

### Data

This study employs the daily reported COVID-19 cases up to May 5^th^, 2021 provided by the Centre for Health Protection (CHP) of the Department of Health in Hong Kong, which are freely accessible on the government’s open-data platform (https://data.gov.hk/en-data/dataset/hk-dh-chpsebcddr-novel-infectious-agent). All recorded cases of infection are laboratory- and clinical-confirmed via nasopharyngeal swab and PCR with reverse transcription (RT-PCR) (Wu et al., 2020). The data depicts the details of the confirmed cases that are related to individual patients, comprising case number, reported and onset dates, case classification, and other demographic information. Such empirical evidence in Hong Kong can provide an insight into studying the process of case spread.

In response to the imported COVID-19 cases, the government has fully implemented a mandatory quarantine scheme on December 25, 2020 on all inbound visitors, aiming to cut off the contact between asymptomatic cases and local communities. We assume that the imported cases are not attributed to the local spread because of the screening and mandatory quarantines imposed on all incoming travelers since December 2021. To estimate the local spread of COVID-19 in Hong Kong, we consider local cases and cases epidemiologically linked to local cases from December 25, 2020 through May 5, 2021.

### Estimation of SEIR model parameter

A classic stochastic SEIR model is proposed to estimate the course of the epidemic development by fitting the observed cumulative daily reported cases in Hong Kong. As shown below, the total population (N) is partitioned into four compartments, which are susceptible (S), exposed (E), infectious (I), and removed/recovered (R). Among these compartments, removed/recovered (R) is further subdivided into population removed from infectiousness (I) and vaccinated population from susceptible (S) during the period of vaccination.

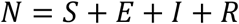

For each time epoch (day), four ordinary differential equations are performed to express the dynamic progression (daily changes) between different epidemiological compartments in the SEIR model, as follows:

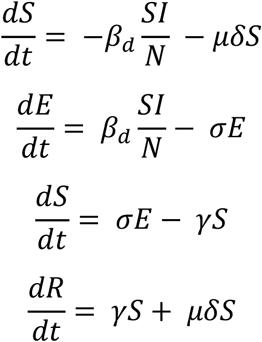

The number of new infections on a daily basis is calculated based on the total contacts between susceptible and infected individuals, of which the transmission rate *β* varies across different regions and countries. The initial *β* is estimated using the basic reproduction number of observed cumulative cases, divided by the infectious period from symptoms onset to the first medical treatment. Here, the incubation period 1/*σ* and infectious period 1/*γ* to be 5.5 days (95% CI: 4.5– 5.8) and 7.1 days (95% CI: 2.5–11.6), respectively (Lauer et al., 2020; Wölfel et al., 2020).

Two modifications are proposed in our model fitting to adapt the local disease etiology. One modification is to consider six main interventions released in the past few months in Hong Kong, including school arrangement, suspension of large events, public facility arrangement, restricting social gathering, using the voluntary check-in system, and compulsory testing, according to a series of NPIs implemented in communities to reduce contacts between the susceptible and potentially infected individuals. The strictness of a specific NPI is represented by using a multiplier to amplify its base value As these interventions have been updated over time, transmission rate β is then multiplied by the coefficient of various implemented NPIs *α*_*NPIS*_, for recapitulating the trend of local infections more precisely, as follows:

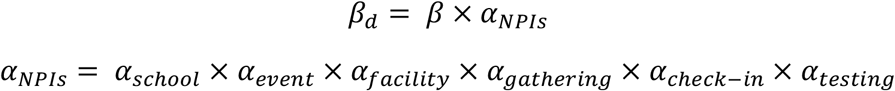

Another modification is the territory-wide COVID-19 vaccination program for all Hong Kong residents since February 27, 2021, which is proposed as an essential public health barrier against COVID-19. According to the cumulative vaccinated population, the Levenberg-Marquardt algorithm is employed to estimate the daily vaccination coverage δ of the susceptible population before May 5, 2021. We further consider the probability of inducing an immune response regarding the vaccinated population, since some individuals are unable to be vaccinated for vaccine breakthrough infections (Hacisuleyman et al., 2021). Therefore, the effectiveness μ of the vaccines adopted in Hong Kong is deployed to specify the vaccinated population that would actually acquire immunity (Baden et al., 2021; Nupus, 2020). The vaccinated population would not infect or be infected by infectious individuals.

All designated parameters are subject to statistical inference and estimated from the daily cumulative reported cases in Hong Kong. The statistical inference is calibrated within a Bayesian optimization framework prior to the Markov Chain Monte Carlo simulation.

### Simulation of the effectiveness of digital contact tracing

Because of the potential virus carriers during the border-reopening phase, it is expected that the COVID-19 resurgence in Hong Kong is highly possible. Therefore, we simulate three baseline scenarios with different vaccination coverages (30%, 50%, and 70%) to understand whether digital contact tracing can substitute the mandatory quarantine while minimizing the importation risk of COVID-19. The model imposes digital contact tracing on all inbound travelers during their entire stay in Hong Kong. Specifically, susceptible local population (S) who closely contact with infectious travelers (I) will be immediately quarantined or isolated, becoming removed population (R). In our simulation, all close contacts are ideally (100%) self-isolated or mandatory quarantined.

Since Hong Kong has imposed extremely strict screening measures including mandatory pre-departure and arrival tests on all inbound travelers, we assume that only susceptible *T*_*susceptiple*_ and asymptomatic travelers *T*_*exposed*_ can enter this city. Hence, the overall importation risk ε is represented as the percentage of inbound travelers exposed to the virus upon arrival based on the total imported cases and total arrival population during the period of observation. Such a setting can simulate a more realistic travel behavior, as most people tend to visit a city for a few days. Thus, the daily inbound population exposed to the virus is represented as a sum of a random sample drawn from a Poisson distribution, as follows:

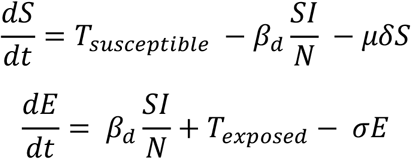

As suggested by the total historical arrivals in 2019 with an average stay of 3.3 nights (Tourism Commission, 2020), we modify our model by deducting a daily inflow population of 152,473 from our model based on samples from a Poisson distribution (λ = 3.3). Therefore, the total population exiting Hong Kong *EX*_*d*_ on day *d* is represented as follows:

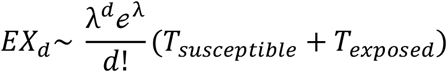

### Incorporating joint inflow population control and community NPIs

Although digital contact tracing can help contain the local infection, its implementation cannot ensure a promising health outcome in the most realistic or the worst-case scenario (30% vaccination coverage). Considering that only a small susceptible population is vaccinated, in addition to contact tracing, it is imperative to estimate the joint effects of controlling arrival population and implementing community NPIs (Brauner et al., 2021). In this regard, a set of scenarios combining the daily quota and the community NPIs are proposed.

With respect to the daily quota, three different border reopening policies are simulated. Compared to the baseline scenario with a daily arrival population of 152,473, two other quotas are set: 76,236 per day (50% of historical arrivals) and 10,000 per day, respectively. Limiting the daily inbound population can control the potential importation of virus carriers, thus reducing the number of active exposure in the SEIR model. Simultaneously, community NPIs are also implemented during the border-reopening phase, with the aim to lower the transmission rate between the infectious and susceptible populations. The proposed community NPIs are on a moderate-and-strong level with school arrangement, suspension of large events, public facility arrangement, restricting social gathering, using the voluntary check-in system, and compulsory testing (Supplementary Table 1). Unlike controlling for the arrival population, NPIs used in our simulations encourage people to practice social and physical distancing. However, compared to mandatory contact tracing and restricting arrival population, community NPIs have severe socioeconomic adversities in people’s daily life. Thus, the NPIs adopted in this study exclude the most restrictive scenarios, such as regional lockdown and school closures.

## Data Availability

The dataset and codes for the this work will be available in Github

## Supplementary

**Table 1.** Description of non-pharmaceutical interventions (NPIs) implemented in Hong Kong from December 2020 to January 2021

| NPI | Level | Description |
| --- | --- | --- |
| School arrangement | Strict | Suspension of face-to-face classes for all schools |
|  | Moderate | Face-to-face classes with the number of students capped at one-third of the total number of students. |
|  | Mild | Face-to-face classes with the number of students capped at one-third of the total number of students. |
| Suspension of large events | Strict | Large-scale events are canceled to reduce public risk |
|  | Moderate | Large-scale events are partially opened |
| Public facility arrangement | Strict | Public facilities are temporarily closed (cultural, recreation and sports activities) |
|  | Moderate | Public facilities are partially opened |
|  | Mild | Public facilities are opened with restrictions |
| Restricting social gathering | Strict | No more than two persons are allowed to gather within any public space |
|  | Moderate | No more than four persons are allowed to gather within any public space |
| Voluntary check-in system |  | A mobile app allowing users to record their whereabouts and get exposure notification. |
| Compulsory testing |  | Restricted districts are delineated based on confirmed case number with a "restriction-testing declaration" to require all residents for a virus testing. |

**Table 2.**
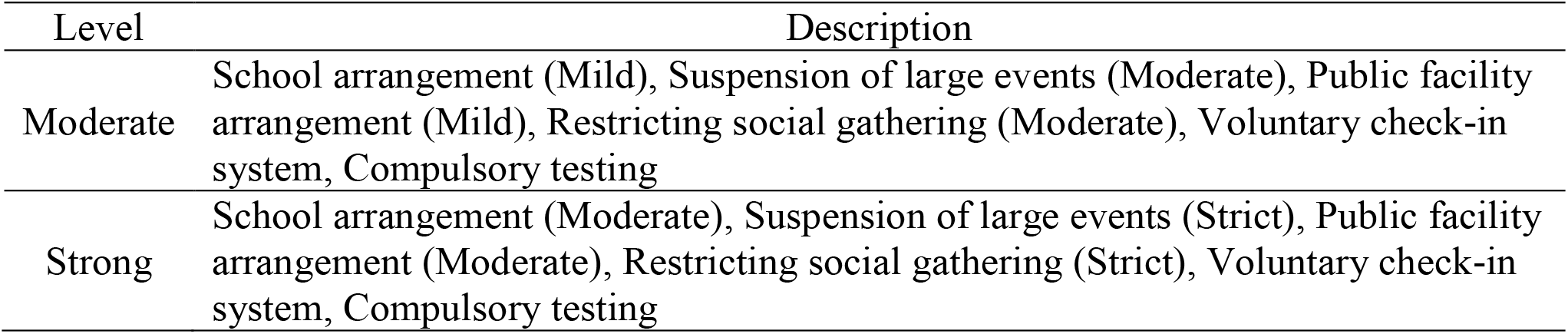
Description of combined community NPIs implemented in designated scenarios

**Table 3.** Implementing a daily quota for inbound travelers and community NPIs with contact tracing (100% successful for individuals) in containing importation risk for reopening borders.

| Daily quota | Community NPIs | Cumulative cases on the 60<sup>th</sup> days (95%CI) | Cumulative cases on the 60<sup>th</sup> days (95%CI) | Reduction rate of cases (%) |
| --- | --- | --- | --- | --- |
|  |  | No contact tracing | Contact tracing |  |
| No | No | 132243(41353-220293) | 10429(8763-12447) | 92.11 |
|  | Moderate | 18362(15678-21544) | 9992(9213-11150) | 45.58 |
|  | Strong | 15568(13790-17658) | 10061(9168-11119) | 35.37 |
| 50% of historical inflow | No | 60598(18690-98721) | 5008(3859-7105) | 91.73 |
|  | Moderate | 8994(7521-10446) | 5057(4490-5592) | 43.77 |
|  | Strong | 7599(6706-8590) | 5039(4520-5549) | 33.68 |
| 10,000 | No | 5618(1063-11676) | 978(151-2069) | 82.59 |
|  | Moderate | 995(753-1222) | 62(437-699) | 43.51 |
|  | Strong | 802(654-1006) | 549(440-697) | 31.54 |

Note: The reduction rate refers to the percentage of cases that could be reduced if the digital contact tracing is applied, compared with the identical scenario without contact tracing.

**Table 4.** Implementing a daily quota for inbound travelers and community NPIs with contact tracing (90% successful for individuals) in containing importation risk for reopening borders.

| Daily quota | Community NPIs | Cumulative cases on the 60<sup>th</sup> days (95%CI) | Cumulative cases on the 60<sup>th</sup> days (95%CI) | Reduction rate of cases (%) |
| --- | --- | --- | --- | --- |
|  |  | No contact tracing | Contact tracing |  |
| No | No | 129638(32387-229935) | 20723(12288-28314) | 84.01 |
|  | Moderate | 18460(15548-21643) | 10983(9913-12042) | 40.50 |
|  | Strong | 15588(13907-17489) | 10705(9683-11670) | 31.32 |
| 50% of historical inflow | No | 57278(17796-98606) | 10571(5496-14700) | 81.54 |
|  | Moderate | 8772(7613-10354) | 5473(4792-5978) | 37.60 |
|  | Strong | 7762(6739-8642) | 5208(4711-5859) | 32.90 |
| 10,000 | No | 6700(2166-10102) | 1113(306-2687) | 83.38 |
|  | Moderate | 960(725-1210) | 655(469-733) | 31.77 |
|  | Strong | 859(647-1027) | 634(457-716) | 26.19 |

Note: The reduction rate refers to the percentage of cases that could be reduced if the digital contact tracing is applied, compared with the identical scenario without contact tracing.

**Fig. 1.**
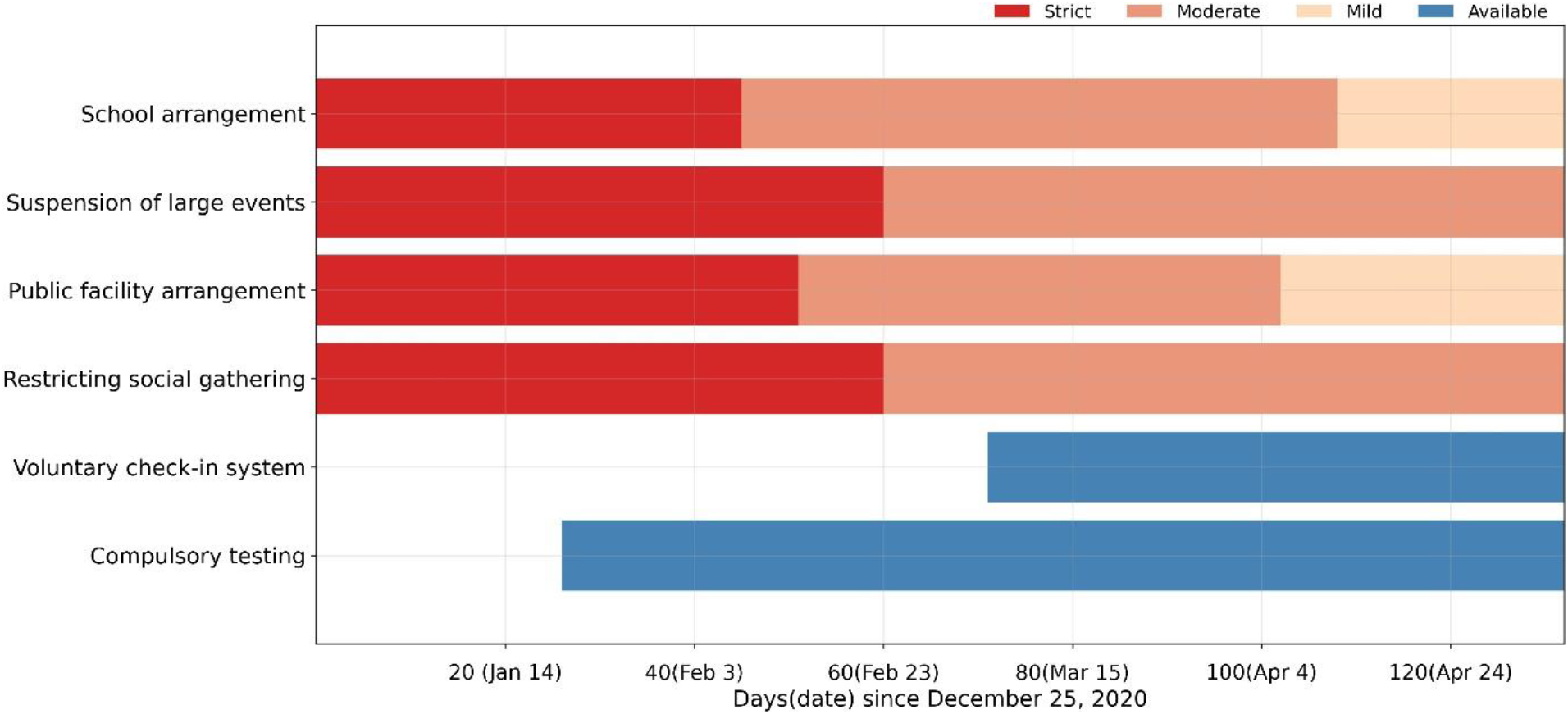
Gantt chart for the implementation period of NPIs. Different colors represent levels of strictness in terms of the implemented NPIs.

**Fig. 2.**
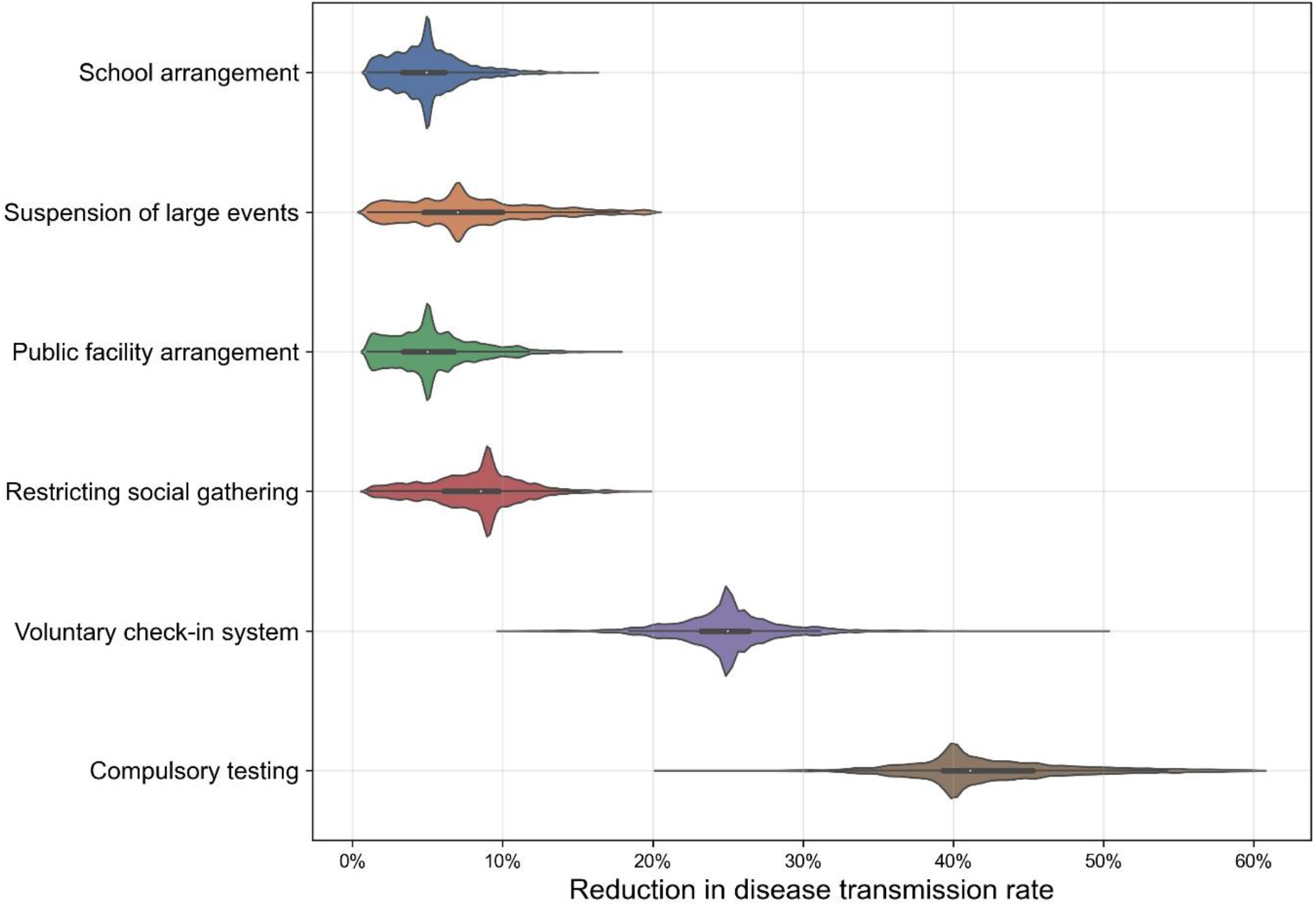
Individual NPI effectiveness in reducing disease transmissivity deriving from the modified model fitting with observed daily reported cases. Results are shown using a violin plot and the box plot inside each violin display median and interquartile range.

**Fig. 3.**
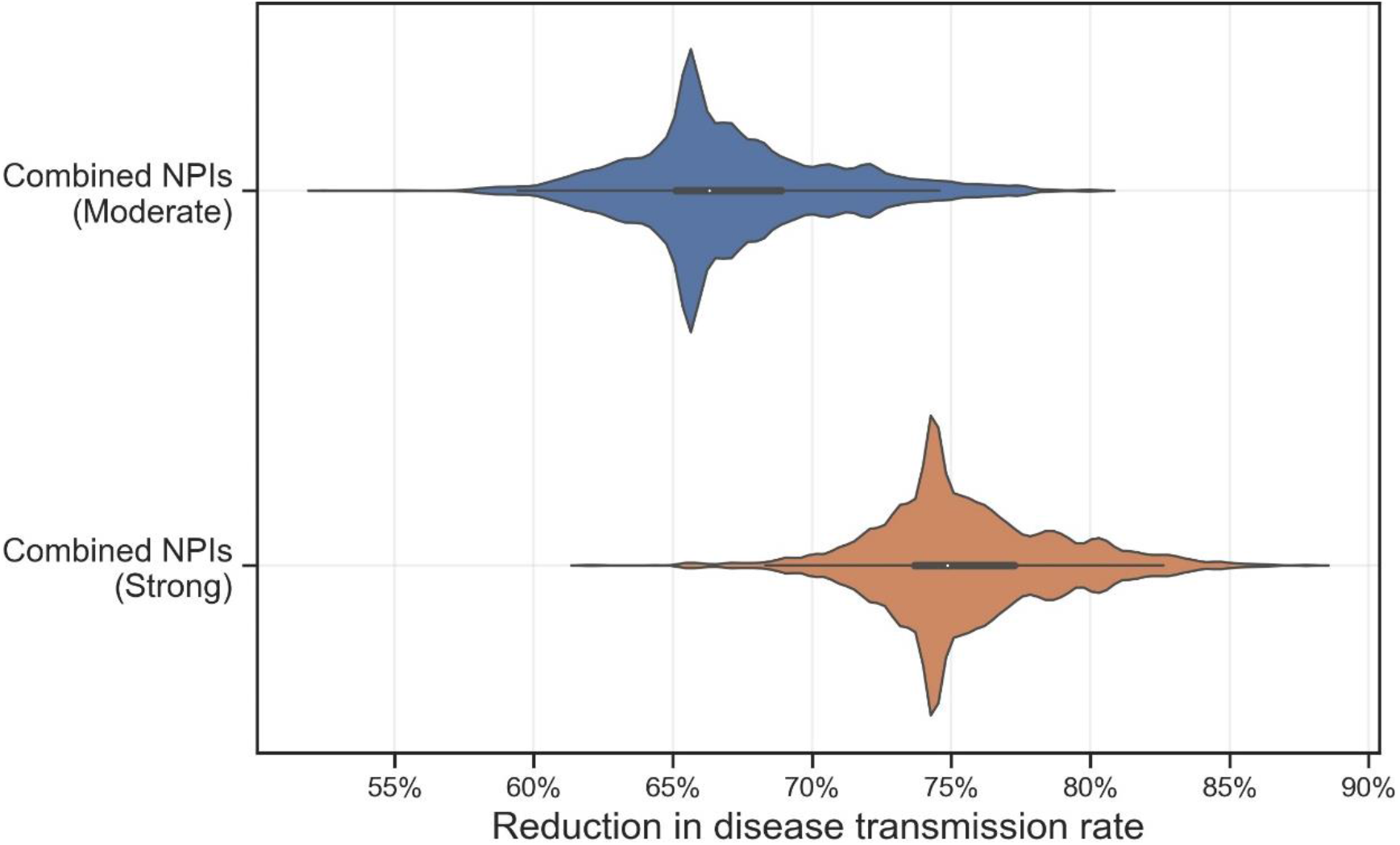
Combined NPI effectiveness in reducing disease transmissivity deriving from the modified model fitting with observed daily reported cases. Results are shown using a violin plot and the box plot inside each violin display median and interquartile range.

**Fig. 4.**
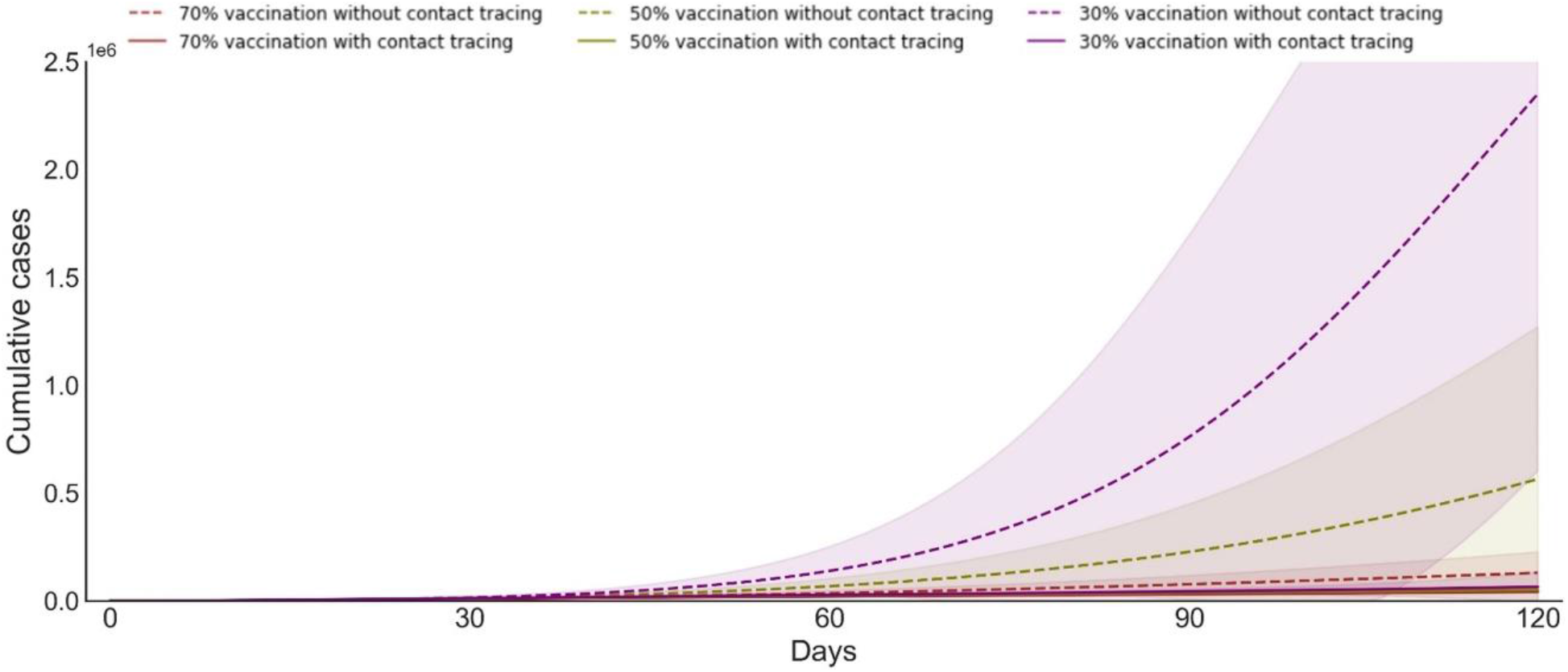
Estimated cumulative daily cases under three different vaccination coverage. The shaded area shows the 95% CI of estimated cumulative cases. Solid and dotted lines represent the scenarios with and without contact tracing, respectively. Different colors represent scenarios with 70%, 50%, and 30% vaccination coverage.

**Fig. 5.**
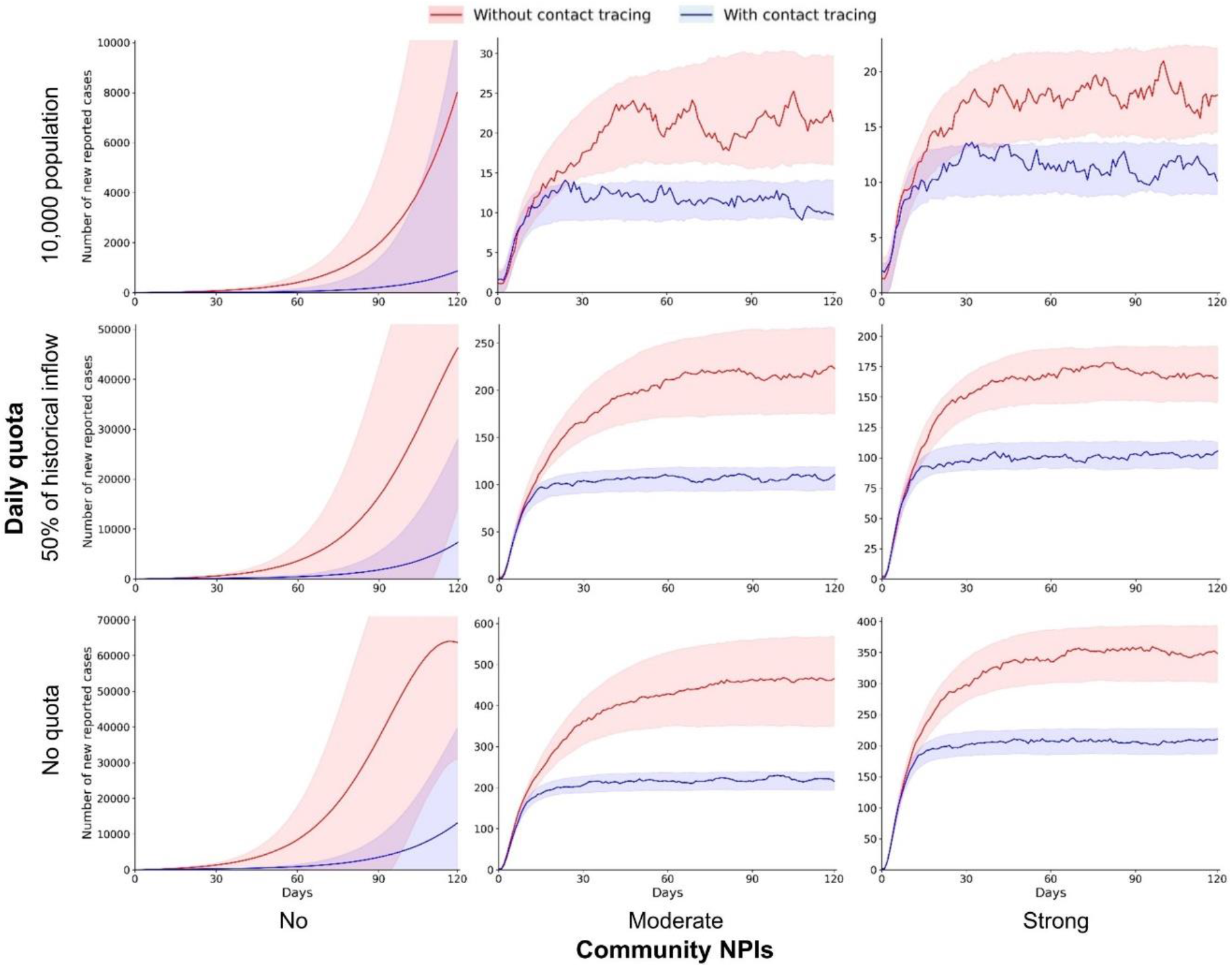
Implementing a daily quota for inbound travelers and community NPIs to contain the importation risk during the 120-day simulation period (90% successful using contact tracing for individuals). The number of daily reported cases are estimated under scenarios with different daily quotas (no quota, 50% of historical inflow, and 10,000 population) and community NPIs (no, moderate, and strong). The shaded areas are the 95% CI of estimated daily reported cases.

